# Limitations of molecular and antigen test performance for SARS-CoV-2 in symptomatic and asymptomatic COVID-19 contacts

**DOI:** 10.1101/2022.02.05.22270481

**Authors:** Matthew L. Robinson, Agha Mirza, Nicholas Gallagher, Alec Boudreau, Lydia Garcia, Tong Yu, Julie Norton, Chun Huai Luo, Abigail Conte, Ruifeng Zhou, Kim Kafka, Justin Hardick, David D. McManus, Laura L. Gibson, Andrew Pekosz, Heba Mostafa, Yukari C. Manabe

**Affiliations:** Division of Infectious Diseases, Department of Medicine, Johns Hopkins University School of Medicine, Baltimore, MD, USA; Department of Pathology, Johns Hopkins University School of Medicine, Baltimore, MD, USA; Department of Molecular Microbiology and Immunology, Johns Hopkins Bloomberg School of Public Health, Baltimore, MD, USA; Clinical Research Unit, Johns Hopkins Institute for Clinical and Translational Research Baltimore, MD USA; Department of Medicine, University of Massachusetts T.H. Chan Medical School, Worcester, MA USA; Division of Infectious Diseases and Immunology, Departments of Medicine and Pediatrics, University of Massachusetts Medical School, Worcester, MA, USA

## Abstract

**Objectives:** COVID-19 has brought unprecedented attention to the crucial role of diagnostics in pandemic control. We compared SARS-CoV-2 test performance by sample type and modality in close contacts of SARS-CoV-2 cases.

**Methods:** Close contacts of SARS-CoV-2 positive individuals were enrolled after informed consent. Clinician-collected nasopharyngeal (NP) swabs in viral transport media (VTM) were tested with a nucleic acid test (NAT). NP VTM and self-collected passive drool were tested using the PerkinElmer real-time reverse transcription PCR (RT-PCR) assay. For the first 4 months of study, mid-turbinate swabs were tested using the BD Veritor rapid antigen test. NAT positive NP samples were tested for infectivity using a VeroE6TMPRSS2 cell culture model.

**Results:** Between November 17, 2020, and October 1, 2021, 235 close contacts of SARS-CoV-2 cases were recruited, including 95 with symptoms (82% symptomatic for ≤5 days) and 140 asymptomatic individuals. NP swab reference tests were positive for 53 (22.6%) participants; 24/50 (48%) were culture positive. PerkinElmer testing of NP and saliva samples identified an additional 28 (11.9%) SARS-CoV-2 cases who tested negative by clinical NAT. Antigen tests performed for 99 close contacts showed 83% positive percent agreement (PPA) with reference NAT among early symptomatic persons, but 18% PPA in others; antigen tests in 8 of 11 (72.7%) culture-positive participants were positive.

**Conclusions:** Contacts of SARS-CoV-2 cases may be falsely negative early after contact, which more sensitive platforms may identify. Repeat or serial SARS-CoV-2 testing with both antigen and molecular assays may be warranted for individuals with high pretest probability for infection.

## INTRODUCTION

As global COVID-19 cases exceed 250 million by December, 2021,(1) SARS-CoV-2 diagnosis remains a critically important public health priority for individuals to know their status, and, from a public health standpoint, to understand the amount of circulating virus and risk of infection. Sustained demand for SARS-CoV-2 testing has accelerated the development and deployment of multiple competing testing technologies, sample types, and approaches.(2, 3) Rapid diagnostic tests that use a lateral flow assay (LFA) to detect SARS-CoV-2 antigen offer scale, convenience, and potential for home use but suffer from reduced sensitivity compared to nucleic acid tests (NAT).(4)

In early symptomatic disease - within 5 days of symptom onset - infected persons are likely to have high viral loads such that both antigen and molecular tests are likely to perform well. For individuals who are close contacts,(5) the viral burden of the index case has been associated with risk of transmission.(6) However, in the pre-symptomatic phase when viral burdens are still low,(7) false negative antigen tests can occur. A recent study from the Netherlands in close contacts showed that only 64% of SARS-CoV-2 molecular test positive close contacts were identified by antigen testing; antigen test sensitivity was as low as 58.7% and 84.2% in symptomatic individuals.(8)

We sought to evaluate the performance of molecular and antigen testing in symptomatic and asymptomatic close contacts of SARS-CoV-2 confirmed individuals. We also sought to assess the sensitivity of both molecular and antigen tests in nasal swabs and molecular tests in saliva to diagnose close contacts with infectious virus in NP swab viral transport media (VTM) using the VeroE6TMPRS2 cell model.

## METHODS

### Patient population and study design

Participants were recruited through posters, online advertisement, and social media written in English and Spanish. Direct outreach targeted close contacts of persons who tested positive for SARS-CoV-2. Under a partial waiver of HIPAA authorization, study staff called patients who tested positive for SARS-CoV-2 at any Johns Hopkins Medical System site to provide study information to their close contacts, defined as individuals who spent >15 minutes at <6 feet in the 5 days after symptom onset or test positivity.(9) Interested close contacts were able to participate by contacting a dedicated study phone number. Study staff obtained informed consent from willing and eligible participants using a script administered either over the phone or in person. Participants were scheduled for same- or next-day testing at one of two outdoor outpatient testing sites. Two weeks after specimen collection, the study staff called the participant to administer a brief questionnaire which assessed ongoing or new COVID-19 symptoms, hospitalization and any other SARS-CoV-2 test results that the participant may have obtained after enrollment. The study was approved by the Johns Hopkins School of Medicine Internal Review Board.

### Sample collection

NP swabs were collected by a trained nurse on all participants and immediately inoculated into VTM. Study staff additionally observed self-collection of a mid-turbinate swab for antigen testing, and 2 mL of passive drool saliva in a sterile urine cup. All specimens were transported on ice to the clinical laboratory using a medical courier service. The transit time from sample collection sites to the laboratory was 15-20 minutes, and samples were processed immediately upon receipt.

### Reference standard nucleic acid testing

All molecular testing took place in the Johns Hopkins Molecular Virology Laboratory, which is Clinical Laboratory Improvement Amendments of 1988 (CLIA) certified. For each assay, testing was performed per manufacturer Food and Drug Administration emergency use authorization (EUA) instructions: NeuMoDx™ SARS-CoV-2 assay (NeuMoDx, Ann Arbor, Michigan)(10), Roche, RealStar® SARS-CoV-2 RT-PCR (Altona Diagnostics, Hamburg, Germany), Hologic Aptima® SARS-CoV-2 assay (Hologic, Bedford, MA),(11) PerkinElmer New Coronavirus Nucleic Acid Detection Kit (PerkinElmer, Inc. Austin, Texas),(12) BD Veritor SARS-CoV-2 (Becton Dickinson, Sparks, Maryland) and interpreted using the Veritor Plus Analyzer.(13) The reference clinical assay performed on fresh NP swab VTM was either the NeuMoDx, Roche, Aptima, Cepheid Xpert, or Altona assays. Remnant specimen was aliquoted and stored frozen at -80^°^C and then 300 µl thawed for extraction followed by RT-PCR on the PerkinElmer instrument. Similarly, fresh saliva was aliquoted, frozen, and then in batches 300 µl of saliva was processed using the same methods. BD Veritor testing occurred within 6 hours of collection with all mid-turbinate swabs stored on ice. At the time of test performance and interpretation, staff were blinded to results of other testing modalities and sample types. Antigen testing was performed in a separate laboratory space independent of molecular test facilities.

### Virus culture from nasal swabs

VeroE6TMPRSS2 cells were grown in complete medium (CM) consisting of DMEM with 10% fetal bovine serum (Gibco), 1 mM glutamine (Invitrogen), 1 mM sodium pyruvate (Invitrogen), 100 U/ml of penicillin (Invitrogen), and 100 μg/ml of streptomycin (Invitrogen).(14) Viral infectivity was assessed on VeroE6TMPRSS2 cells as previously described using infection media (IM; identical to CM except the FBS is reduced to 2.5%).(15) When a cytopathic effect was visible in >50% of cells in a given well, the supernatant was harvested. The presence of SARS-CoV-2 was confirmed through RTqPCR as described previously by extracting RNA from the cell culture supernatant using the Qiagen viral RNA isolation kit and performing RTqPCR using the N1 and N2 SARS-CoV-2-specific primers and probes in addition to primers and probes for human RNaseP gene using synthetic RNA target sequences to establish a standard curve.(16)

### Statistical analysis

Clinical characteristics of participants with positive clinical reference test results were compared to participants with negative results. Categorical variables were compared using the chi-squared test or exact Fisher test for comparisons including groups with <5 expected frequencies. Continuous variables were compared using the Kruskal-Wallis test. As SARS-CoV-2 molecular tests are highly specific, in cases of discordant molecular results by modality (standard clinical reference test or PerkinElmer RT-PCR) or sample type (NP or saliva), participants who tested positive by any molecular test were considered to have SARS-CoV-2. Simple frequencies were calculated for the proportion of infectious samples (as measured by cell culture) that were positive by antigen and molecular testing. Antigen test performance characteristics were reported as positive and negative percent agreement with the clinical reference test and calculated as simple proportions with two-sided 95% confidence intervals (CI).

## RESULTS

### Clinical Characteristics among all participants, by SARS-CoV-2 diagnosis

Between November 17, 2020, and October 1, 2021, 327 participants were consented, among whom 301 presented for sample collection and provided information to evaluate their status as a close contact of a laboratory-confirmed SARS-CoV-2 case (Figure 1). There were 235 close contacts (upon presentation, n=66 were deemed not to meet close contact criteria, Figure 1) who comprise the analyzed cohort among whom 95 were symptomatic and 140 were asymptomatic. Of the symptomatic contacts, 82% (78/95) had symptoms for ≤5 days. Table 1 shows the baseline characteristics of the participants. Overall, the median age was 38.0 (IQR 29.0-50.5), 52.3% male. There were 148 (63%) participants who reported having healthcare coverage. The standard clinical reference in NP samples was positive for 53 (23%) participants. Those who were SARS-CoV-2 test positive compared to those who were negative were more likely to be unvaccinated (83.0% vs 71.4%, p = 0.050), Black (22.6% vs 15.9%) and of Hispanic ethnicity (54.7% vs 39.6%). Among the cohort of COVID-19 contacts, 32.1% of those who were test positive were asymptomatic, which may include contacts in the early, pre-symptomatic phase or asymptomatic infections. The most common symptoms among the NP swab SARS-CoV-2 positive patients were cough (41.5%), runny nose (41.5%), muscle aches (35.8%), fever or chills (32.1%), and scratchy throat (26.4%).

**Figure 1:**
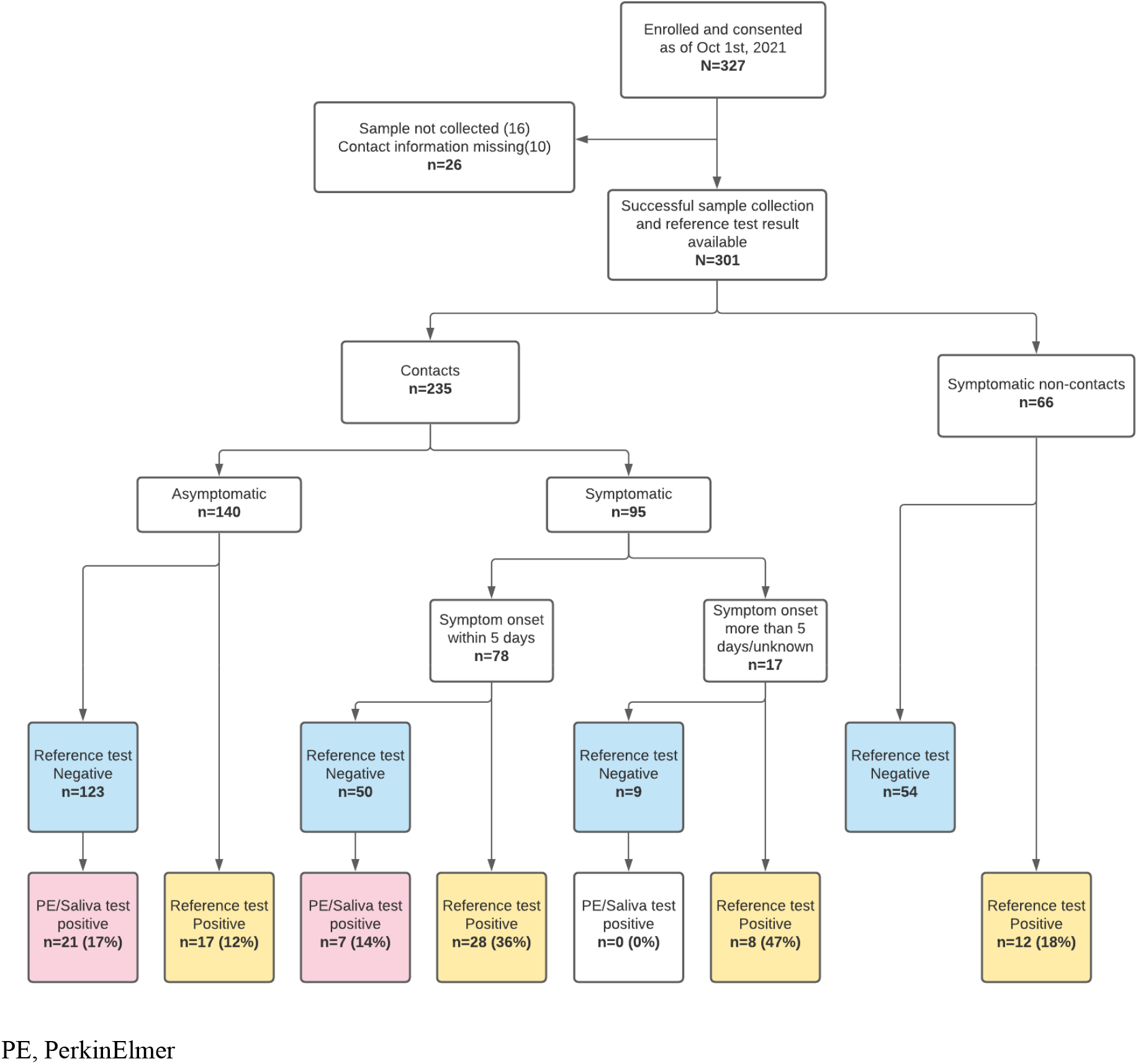
Participants enrolled and molecular test results.

**Table 1:**
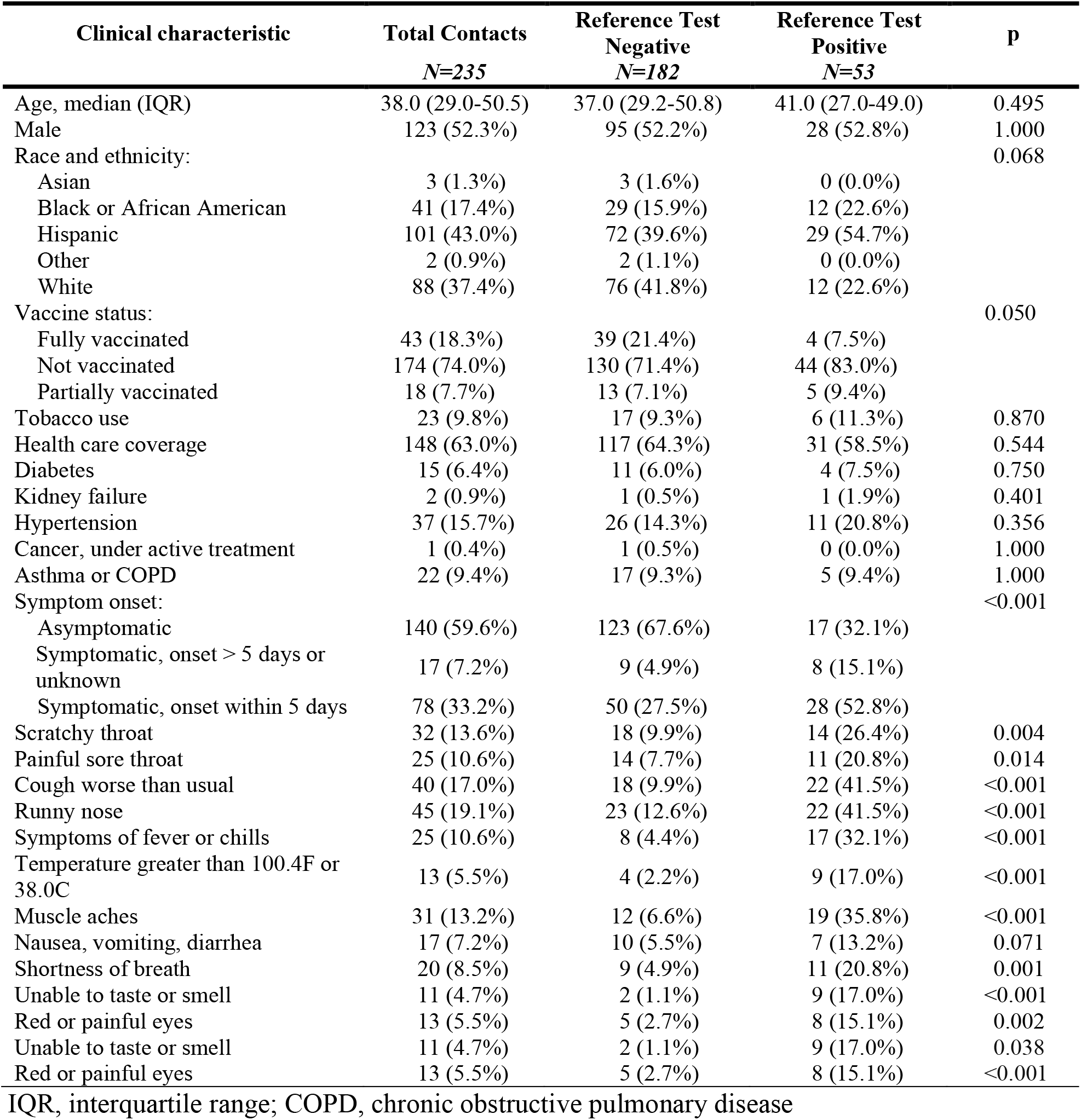
Clinical characteristics among close contacts stratified by SARS-CoV-2 reference test result.

### Molecular test and viral culture results

The NP swab reference NAT was positive for 22.6% (53/235) of participants who were close contacts of SARS-CoV-2 cases (Table 2). Among 50 of 53 reference positive samples which underwent viral culture, 25 (50%) were culture positive. Remnant frozen NP swab VTM for 158 of the 182 SARS-CoV-2 close contacts who tested negative by the reference test was retested by PerkinElmer RT-PCR and positive for 13 (8%) samples with a median cycle threshold (Ct) of 36.8 (IQR 36.0-38.6). Saliva testing using PerkinElmer RT-PCR was performed for 186 (79%) close contacts, including 143 of 182 (79%) reference test negative participants. Saliva was positive for 37 of 43 (86%) reference positive participants and 17 of 143 (12%) reference negative participants. Combined, PerkinElmer RT-PCR of NP and saliva samples identified SARS-CoV-2 in an additional 21 asymptomatic close contacts and 7 symptomatic close contacts not identified by the NP reference test. The median Ct value of symptomatic close contacts was lower than asymptomatic close contacts for both NP (22.0, IQR 17.7-28.5 vs 34.8, IQR 31.4-36.9) and saliva (25.8, IQR 22.5-31.5 vs 33.4, IQR 26.5-36.7) samples. Among the 67 participants that tested positive by any NAT and had a saliva NAT result, the sensitivity of saliva NAT was 80.6% and NP was 77.6% (Table S1). Among symptomatic patients, patients with systemic symptoms of fever and muscle ache were more likely to test positive by both NP and saliva samples and patients with runny nose were more likely to test positive by NP only.

**Table 2:**
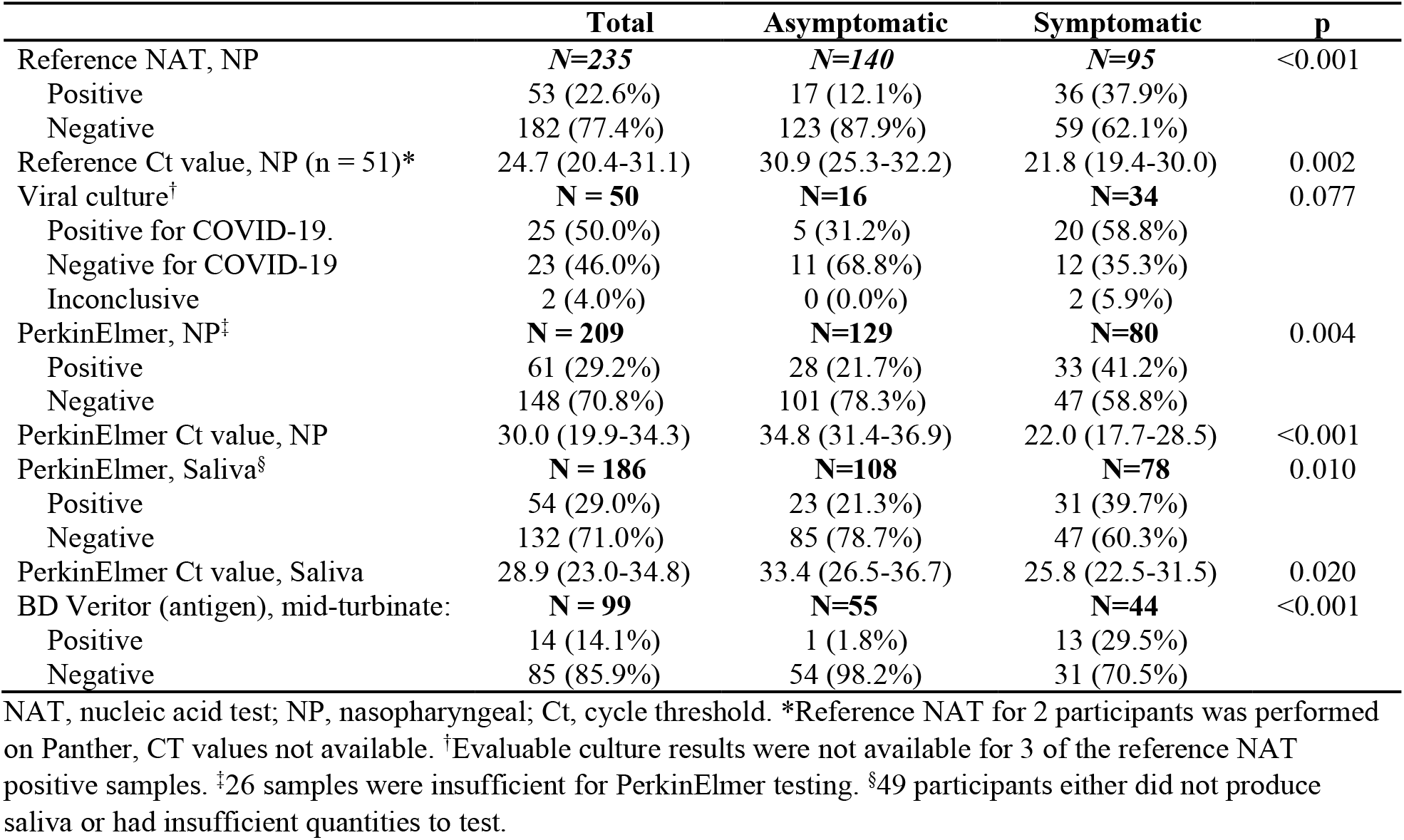
SARS-CoV-2 testing among contacts by presence of symptoms.

|  | **Total** | **Asymptomatic** | **Symptomatic** | **p** |
| --- | --- | --- | --- | --- |
| Reference NAT, NP | **N=235** | **N=140** | **N=95** | <0.001 |
| Positive | 53 (22.6%) | 17 (12.1%) | 36 (37.9%) |  |
| Negative | 182 (77.4%) | 123 (87.9%) | 59 (62.1%) |  |
| Reference Ct value, NP (n = 51)* | 24.7 (20.4-31.1) | 30.9 (25.3-32.2) | 21.8 (19.4-30.0) | 0.002 |
| Viral culture† | **N = 50** | **N=16** | **N=34** | 0.077 |
| Positive for COVID-19. | 25 (50.0%) | 5 (31.2%) | 20 (58.8%) |  |
| Negative for COVID-19 | 23 (46.0%) | 11 (68.8%) | 12 (35.3%) |  |
| Inconclusive | 2 (4.0%) | 0 (0.0%) | 2 (5.9%) |  |
| PerkinElmer, NP‡ | **N = 209** | **N=129** | **N=80** | 0.004 |
| Positive | 61 (29.2%) | 28 (21.7%) | 33 (41.2%) |  |
| Negative | 148 (70.8%) | 101 (78.3%) | 47 (58.8%) |  |
| PerkinElmer Ct value, NP | 30.0 (19.9-34.3) | 34.8 (31.4-36.9) | 22.0 (17.7-28.5) | <0.001 |
| PerkinElmer, Saliva§ | **N = 186** | **N=108** | **N=78** | 0.010 |
| Positive | 54 (29.0%) | 23 (21.3%) | 31 (39.7%) |  |
| Negative | 132 (71.0%) | 85 (78.7%) | 47 (60.3%) |  |
| PerkinElmer Ct value, Saliva | 28.9 (23.0-34.8) | 33.4 (26.5-36.7) | 25.8 (22.5-31.5) | 0.020 |
| BD Veritor (antigen), mid-turbinate: | **N = 99** | **N=55** | **N=44** | <0.001 |
| Positive | 14 (14.1%) | 1 (1.8%) | 13 (29.5%) |  |
| Negative | 85 (85.9%) | 54 (98.2%) | 31 (70.5%) |  |

NAT, nucleic acid test; NP, nasopharyngeal; Ct, cycle threshold. \*Reference NAT for 2 participants was performed on Panther, CT values not available. †Evaluable culture results were not available for 3 of the reference NAT positive samples. ‡26 samples were insufficient for PerkinElmer testing. §49 participants either did not produce saliva or had insufficient quantities to test.

### Antigen test results

From November 17, 2020, to March 11, 2021, the initial 126 participants underwent antigen testing of a mid-turbinate swab using the BD Veritor lateral flow assay among whom, 99 were close contacts. Among close contacts who underwent antigen testing, 55 (56%) were asymptomatic, 35 (35%) were symptomatic and within the first 5 days of symptom onset, and 9 (9%) were symptomatic but beyond 5 days of symptom onset. Among symptomatic close contacts in the first 5 days of symptom onset (intended use case for the BD Veritor test), the positive percent agreement (PPA) was 83.3% (95% CI 50.9-97.1) and negative percent agreement was 95.7% (95% CI 76.0-99.8) compared to the reference molecular test. However, among asymptomatic persons or persons after 5 days of symptom onset, the PPA was 18.2% (95% CI 3.2-52.2). The median reference test Ct for participants who tested positive by reference test and negative by BD Veritor was 31.6 for asymptomatic participants, 30.8 for symptomatic in the first 5 days of symptom onset, and 30.3 for participants beyond 5 days of symptom onset (Figure 2A). Among the 11 close contacts that were culture positive with BD Veritor results, 8 (72.7%) were positive by BD Veritor (Figure 2B). Among the 3 that were missed, 2 were asymptomatic.

**Figure 2:**
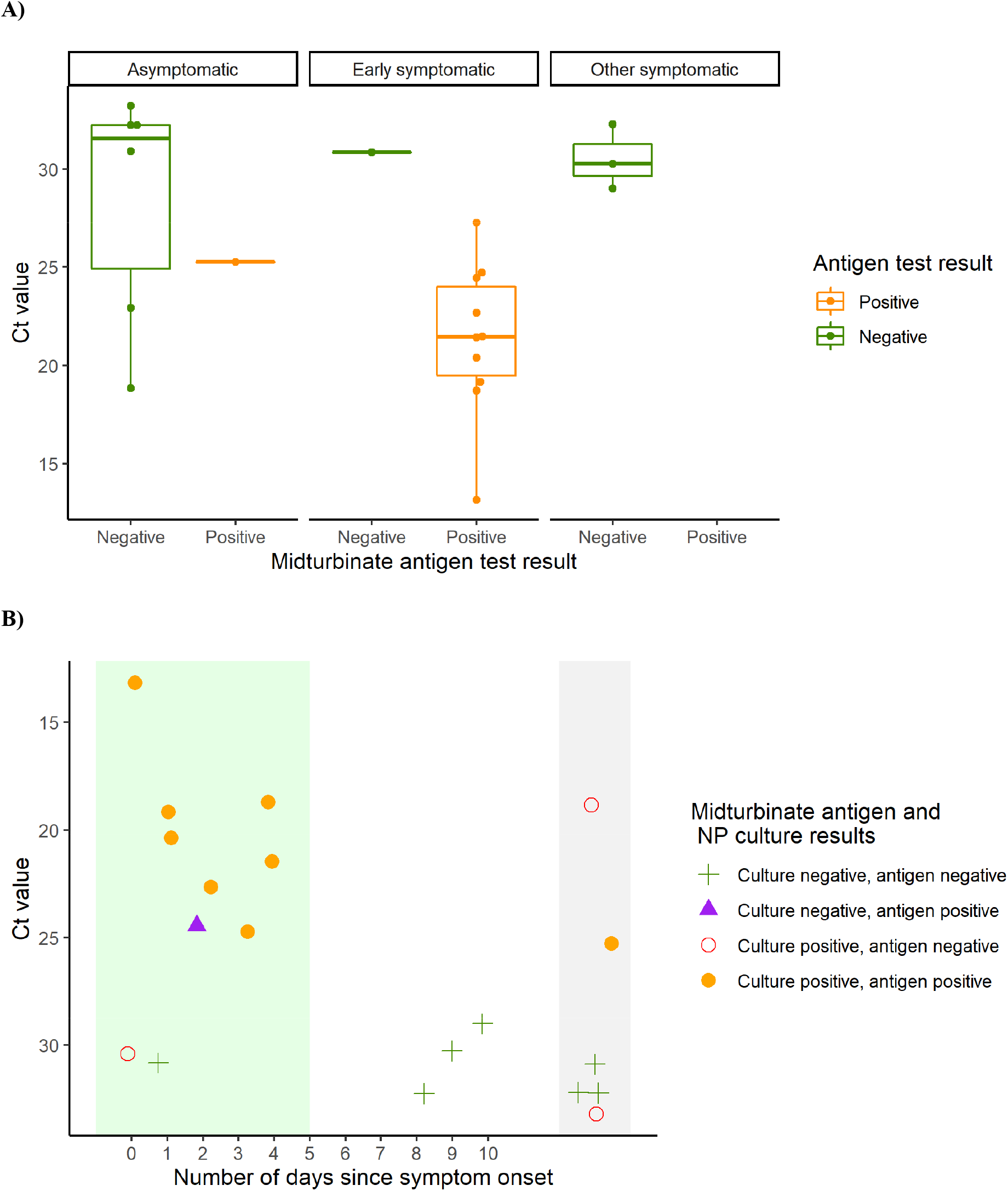
Antigen test positivity of contacts: A) the cycle threshold (Ct) values of the reference assay by stratified by asymptomatic, early symptomatic ≤ 5 days, and symptomatic >5 days participants of the RT-PCR positive by the reference assay for which there were antigen lateral flow results (n=99), and B) Culture, antigen result and Ct of reference assay for all RT-PCR positive NP samples arrayed by early symptomatic (green shaded), symptomatic (white), and asymptomatic (grey)

## DISCUSSION

In this study, we report the relative performance of multiple molecular test approaches in a large number of known SARS-CoV-2 NAT-confirmed case contacts. As expected, a high proportion (22.6%) of these contacts were reference lab NP NAT positive. However, an additional 11.6% were positive when both NP VTM and saliva were tested using a more sensitive platform; more than half of asymptomatic close contacts tested falsely negative using a high-complexity reference lab test. Patients with symptomatic SARS-CoV-2 were less likely to be missed by standard NAT testing and there were no missed cases among patients who were not close contacts. Although prior data has shown that approximately half of patients who eventually test positive for SARS-CoV-2 initially test negative and test performance is worse in the first few days after exposure and before symptom onset,(17, 18) work describing the potential to detect additional cases using more sensitive assays and additional specimen types is limited.

The FDA requires that SARS-CoV-2 NATs demonstrate concordance with existing authorized tests to gain EUA(19) which gives the impression that the clinical performance of all EUA SARS-CoV-2 NAT are approximately equivalent. However, among SARS-CoV-2 NAT with FDA EUA, there is a 100-fold difference between the highest and lowest levels of detection (LOD) reported using comparable FDA reference materials.(20) In this study, testing using the PerkinElmer NAT, which has the lowest reported LOD, identified many cases missed by standard reference tests. Another source of missed positives were samples that tested positive in saliva but not NP samples. Similar to prior work, saliva was not more or less sensitive than NP, but it did identify a population of SARS-CoV-2 cases that was only partially overlapping with those found using NP samples.(21-23) If testing resources were not scarce, a strategy of performing high sensitivity NAT on both saliva and NP specimens may be considered to exclude infection in patients with the highest pretest probability for infection.

Although antigen testing identified the majority of culture-positive contacts, a few patients with culture-positive SARS-CoV-2 were missed. It is expected that antigen testing will miss a proportion of SARS-CoV-2 cases, particularly those with lower viral burden (identified by higher Ct values) particularly among asymptomatic individuals; a recent meta-analysis reported a sensitivity of 58.1% (95% CI 40.2% to 74.1%),(24) and more focused studies including asymptomatic close contacts reported a sensitivity of 59-82% compared to NAT.(25, 26) However, the hope has been that persons with potential to transmit SARS-CoV-2 may be excluded, at least in the moment, by a negative antigen test.(27-29) Used as intended, in the first 5 days of symptom onset, only one patient with culture-positive SARS-CoV-2 tested negative by antigen testing, but two asymptomatic patients who tested negative by antigen tests had positive cultures. Recent data from our group and others has shown that serial testing may be a strategy to overcome the lack of sensitivity in pre-symptomatic patients.(23, 30) Daily antigen testing is now being used by some school systems to forego quarantine after exposure.(31, 32)

False negative SARS-CoV-2 test results for asymptomatic close contacts may provide false reassurance, leading to relaxation of isolation measures and greater onward transmission. Our findings suggest that for asymptomatic close contacts with a high pretest probability for infection, testing at a single time point with a standard reference NAT misses some cases and negative antigen testing does not exclude transmissibility. SARS-CoV-2 exposures that are to household members, indoors, and prolonged pose a higher risk of transmission(33) which may warrant additional vigilance including repeat testing for close contacts with high pretest probability for infection. Newer SARS-CoV-2 variants including delta and omicron have higher capacity for transmission than earlier circulating genotypes.(34, 35) Therefore, the consequences for each false negative test on community transmission may be greater now that currently circulating variants are more transmissible than earlier circulating SARS-CoV-2.

Limitations include the possibility that other commercially available antigen tests may outperform the one chosen for use in this study, although direct comparisons of the Veritor antigen test used in this study has shown similar performance characteristics to others.(25, 36) Similarly, differences in the analytical sensitivity of SARS-CoV-2 test platforms may impact the results if this study were to be replicated using alternative SARS-CoV-2 NAT. Performance of the antigen tests was not performed as intended at the point of collection, which could reduce their sensitivity. Most of the positive cases were from unvaccinated persons prior to circulation of delta and omicron variants – as more vaccinated persons test positive for COVID, new data regarding the relationship between pretest probability of infection, test performance characteristics, SARS-CoV-2 variant, and culture positivity may be required. Data regarding timing of participant exposure to an individual with COVID was not available, which limits the ability to identify subgroups of close contacts with better or worse test performance.

Ongoing SARS-CoV-2 transmission with increasingly contagious variants highlights the continued importance of widespread and accurate testing even as the pandemic approaches its third year. Our findings in a large cohort of close contacts demonstrate the limitations of cross-sectional antigen testing to exclude transmissibility, and standard NAT to diagnose COVID-19. Close contacts of individuals with COVID-19 should isolate and may need to test more than once, especially those with the highest risk exposures such as household contacts. Future work is necessary to improve test performance characteristics or implement serial testing to identify patients with SARS-CoV-2 when they are most likely to have transmissible disease.

## Data Availability

Data produced in the present study are available upon reasonable request to the authors.

## Acknowledgements

We acknowledge Madison Conte and Elena Konstant for their contributions to the processing and storage of specimens used in this study. Becton Dickenson provided Veritor test kits for use in this study, but did not contribute to the writing of this manuscript nor critically review its content.

## Funding

This study was funded by the NIH RADx-Tech program under 3U54HL143541-02S2 and U54EB007958-12S1. The views expressed in this manuscript are those of the authors and do not necessarily represent the views of the National Institute of Biomedical Imaging and Bioengineering; the National Heart, Lung, and Blood Institute; the National Institutes of Health, or the U.S. Department of Health and Human Services. Salary support from the National Institutes of Health U54EB007958-13 (YCM, MLR, JH), AI272201400007C (AP, YCM), UM1AI068613 (YCM), U54HL143541, R01HL141434, R01HL137784, R01HL155343, R61HL158541, R01HL137734 (DDM).

## Declaration of competing interests

Becton Dickenson provided Veritor test kits for use in this study but did not contribute to the writing of this manuscript nor critically review its content. DDM reports consulting and research grants from Bristol-Myers Squibb and Pfizer, consulting and research support from Fitbit, consulting and research support from Flexcon, research grant from Boehringer Ingelheim, consulting from Avania, non-financial research support from Apple Computer, consulting/other support from Heart Rhythm Society. LG is on a scientific advisory board for Moderna on projects unrelated to SARS-CoV-2. HHM reports receipt of research contracts from BioRad, DiaSorin, and Hologic. YCM has received tests from Quanterix, Becton-Dickinson, Ceres, and Hologic for research-related purposes, consults for Abbott on subjects unrelated to SARS-CoV-2, and receives funding support to Johns Hopkins University from miDiagnostics.

## Supplementary materials

**Table S1:** Characteristics of nucleic acid test positive contacts, stratified by sample type positive, among patients with saliva nucleic acid test results

|  | Positive by any<br>NAT<br>*N=67* | NP only<br>positive<br>*N=13* | Saliva and NP<br>positive<br>*N=39* | Saliva only<br>positive<br>*N=15* | p |
| --- | --- | --- | --- | --- | --- |
| Age, median (IQR) | 41.0 (33.5-51.0) | 42.0 (33.0-47.0) | 38.0 (29.0-46.5) | 46.0 (38.5-60.5) | 0.136 |
| Male | 42 (62.7%) | 9 (69.2%) | 23 (59.0%) | 10 (66.7%) | 0.830 |
| Race and ethnicity: |  |  |  |  | 0.165 |
| Black or African-American | 11 (16.4%) | 0 (0.0%) | 9 (23.1%) | 2 (13.3%) |  |
| Hispanic | 37 (55.2%) | 10 (76.9%) | 21 (53.8%) | 6 (40.0%) |  |
| Other | 1 (1.5%) | 0 (0.0%) | 1 (2.6%) | 0 (0.0%) |  |
| White | 18 (26.9%) | 3 (23.1%) | 8 (20.5%) | 7 (46.7%) |  |
| Vaccine status: |  |  |  |  | 0.261 |
| Fully vaccinated | 8 (11.9%) | 0 (0.0%) | 4 (10.3%) | 4 (26.7%) |  |
| Not vaccinated | 51 (76.1%) | 11 (84.6%) | 31 (79.5%) | 9 (60.0%) |  |
| Partially vaccinated | 8 (11.9%) | 2 (15.4%) | 4 (10.3%) | 2 (13.3%) |  |
| Tobacco use | 9 (13.4%) | 0 (0.0%) | 7 (17.9%) | 2 (13.3%) | 0.363 |
| Health care coverage | 37 (55.2%) | 4 (30.8%) | 25 (64.1%) | 8 (53.3%) | 0.110 |
| Diabetes | 4 (6.0%) | 1 (7.7%) | 2 (5.1%) | 1 (6.7%) | 1.000 |
| Kidney failure | 1 (1.5%) | 0 (0.0%) | 1 (2.6%) | 0 (0.0%) | 1.000 |
| Hypertension | 9 (13.4%) | 0 (0.0%) | 7 (17.9%) | 2 (13.3%) | 0.363 |
| Asthma or COPD | 4 (6.0%) | 0 (0.0%) | 4 (10.3%) | 0 (0.0%) | 0.478 |
| Symptom onset: |  |  |  |  | 0.151 |
| Asymptomatic | 31 (46.3%) | 8 (61.5%) | 14 (35.9%) | 9 (60.0%) |  |
| Symptomatic, onset > 5 days or unknown | 6 (9.0%) | 2 (15.4%) | 4 (10.3%) | 0 (0.0%) |  |
| Symptomatic, onset within 5 days | 30 (44.8%) | 3 (23.1%) | 21 (53.8%) | 6 (40.0%) |  |
| Days of symptoms | 2.0 (1.0-4.0) | 5.0 (2.0-9.0) | 3.0 (1.0-4.0) | 1.0 (1.0-1.8) | 0.127 |
| Scratchy throat | 13 (19.4%) | 1 (7.7%) | 10 (25.6%) | 2 (13.3%) | 0.363 |
| Painful sore throat | 12 (17.9%) | 1 (7.7%) | 9 (23.1%) | 2 (13.3%) | 0.559 |
| Cough worse than usual | 19 (28.4%) | 2 (15.4%) | 14 (35.9%) | 3 (20.0%) | 0.341 |
| Runny nose | 19 (28.4%) | 3 (23.1%) | 16 (41.0%) | 0 (0.0%) | 0.004 |
| Symptoms of fever or chills | 16 (23.9%) | 1 (7.7%) | 14 (35.9%) | 1 (6.7%) | 0.032 |
| Temperature > 38.0C | 7 (10.4%) | 0 (0.0%) | 7 (17.9%) | 0 (0.0%) | 0.086 |
| Muscle aches | 18 (26.9%) | 0 (0.0%) | 15 (38.5%) | 3 (20.0%) | 0.016 |
| Nausea, vomiting, diarrhea | 7 (10.4%) | 1 (7.7%) | 5 (12.8%) | 1 (6.7%) | 1.000 |
| Shortness of breath | 9 (13.4%) | 0 (0.0%) | 8 (20.5%) | 1 (6.7%) | 0.156 |
| Unable to taste or smell | 8 (11.9%) | 3 (23.1%) | 5 (12.8%) | 0 (0.0%) | 0.157 |
| Red or painful eyes | 7 (10.4%) | 1 (7.7%) | 6 (15.4%) | 0 (0.0%) | 0.305 |

NAT, nucleic acid test; NP, nasopharyngeal; IQR, interquartile range; COPD, chronic obstructive pulmonary disease

## Notes

### Author Declarations

The study was approved by the Johns Hopkins School of Medicine Internal Review Board.

